# The impact of surgical mask-wearing, contact tracing program, and vaccination on COVID-19 transmission in Taiwan from January 2020 to March 2022: a modelling study

**DOI:** 10.1101/2022.06.06.22276025

**Authors:** Tatiana Filonets, Maxim Solovchuk, Wayne Gao

**Author notes:** (T.F.); (M.S.).

## Abstract

The effectiveness of interventions such as public mask-wearing, contact tracing, and vaccination presents an important lesson for control of the further COVID-19 outbreaks without of whole country lockdowns and the restriction of individual movement. We simulated different scenarios of COVID-19 waves in Taiwan from 2020 to the beginning of March 2022 and considered the following interventions: travel restrictions, quarantine of infected individuals, contact tracing, mask-wearing, vaccination, and mass gathering restrictions. We propose an epidemiological compartmental model modified from the susceptible-exposed-infectious-removed (SEIR) model and derive a formula for the basic reproduction number (R_0_) describing its dependence on all investigated parameters. The simulation results are fitted with the official Taiwanese COVID-19 data. Thus, the results demonstrate that the fast introduction of the interventions and maintaining them at a high level are able the outbreak control without strict lockdowns. By estimation of the R_0_, it was shown that it is necessary to maintain on high implementation level of both non- and pharmaceutical intervention types to control the COVID-19 transmission. Our results can be useful as advice or recommendation for public health policies, and our model can be applied for other epidemiological simulation studies.

## 1. Introduction

From the beginning of 2020 when the worldwide COVID-19 outbreak has begun, Taiwan has one of the most successful of COVID-19 controlling story. The Taiwanese government was prepared for the situation due to the emergency response network for novel infectious disease outbreaks established after the SARS outbreak in 2003 [1]. In 2021, Taiwan had a daily maximum of 534 domestic cases reported on May 26, 2021. At the end of March 2022, Taiwan started to change the COVID-19 strategy from zero cases policy to living with the virus. Some restrictions started to relax from March 2022 since a large proportion of the population is vaccinated and the Omicron variant has less severe symptoms. In the current study, we investigated the transmission of different COVID-19 variants in Taiwan: January–March 2020 (original), May–July 2021 (Alpha), July–December 2021 (Delta), and January–March 2022 (Omicron).

While Taiwan is relatively close to mainland China, the country has an excellent record of eradicating local COVID-19 transmission in the face of continued imported cases since January, 2020. In 2019, when the world was unaware that a pandemic would soon strike, Taiwan actively inquired to the World Health Organization (WHO) about seven atypical pneumonia cases isolated in hospitals in Wuhan via the National Focal Point reporting system mandated by the International Health Regulation (IHR) [2]. At the same time, the country implemented onboard screening of passengers on all flights from Wuhan and was among the first countries to ban travelers from the province [3]. During the wave of COVID-19 in January–March 2020, Taiwan quickly reacted to the situation and began controlling the spread of the virus. Globally, Taiwan was an early implementer of non-pharmaceutical interventions, including entry restrictions, screening (health checks) for travelers, quarantining for travelers and infected individuals, and contact tracing based on collected data [3–5]. On February 6, 2020, Taiwan launched the Name-Based Mask Distribution System for the citizens to purchase a specific number of masks [5]. Taiwan government has facilitated mask-wearing by banning the export of masks and taking measures to prevent price gouging. There was no local transmission in the country for over 250 days from April to December 2020 [6].

In 2020, Taiwan could contain the local spread of the virus despite the imported cases waves [2,7]. Taiwan’s ability to flatten and eventually eliminate local COVID-19 transmission in the face of imported cases demonstrates the effectiveness of its robust outbreak containment measures (Figure 1).

**Figure 1.**
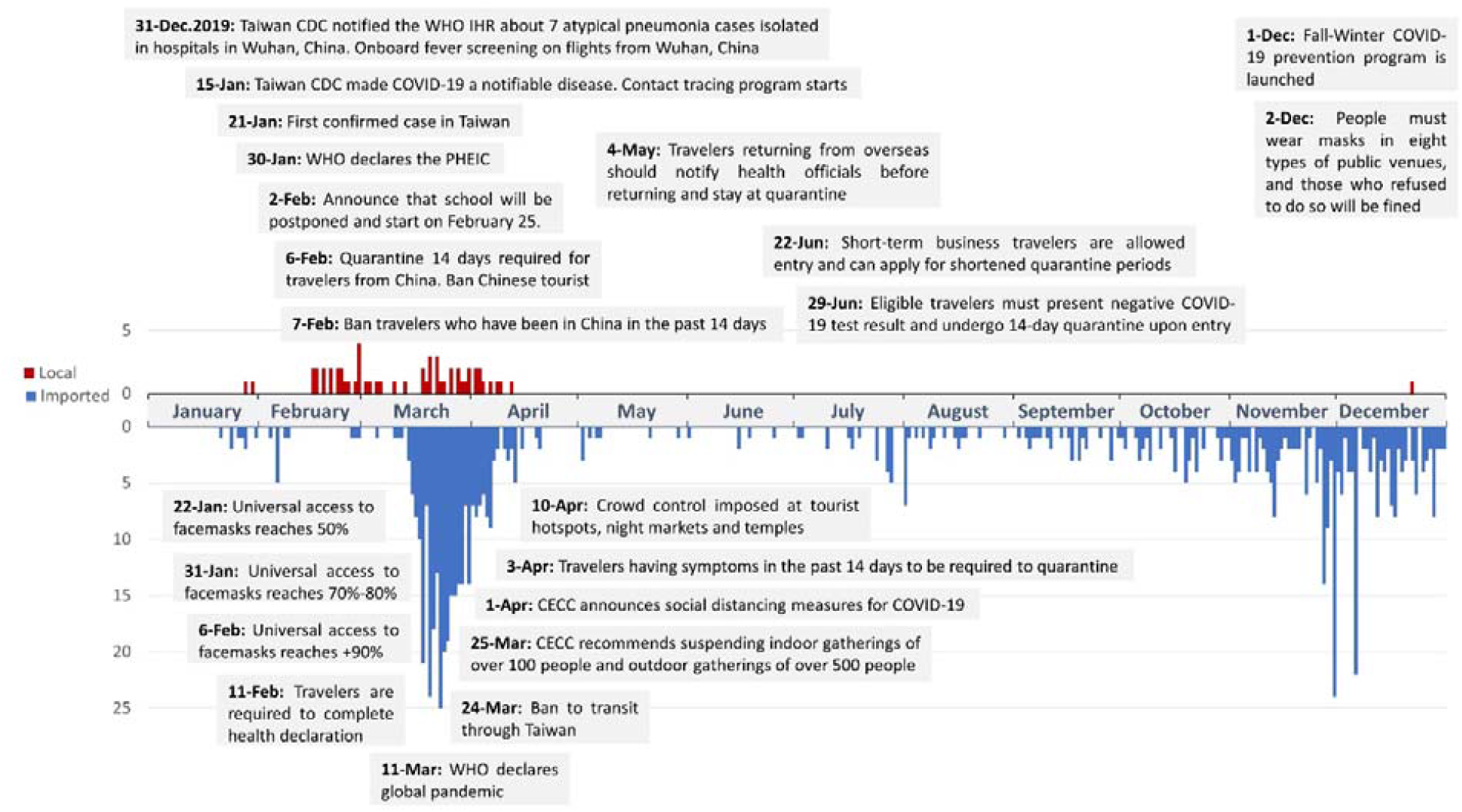
Timeline of officially confirmed COVID-19 cases in 2020 in Taiwan with control measures and their implementation dates.

The government took rapid action to provide a steady supply of face masks to residents. The CECC set the price of masks, used government funds and military personnel to increase mask production [3]. At the end of April 2020, mask production in the country increased from 1.88 million to 19 million units per day [8]. In addition, contact tracing and quarantine requirements have played critical roles in controlling the spread of COVID-19. Taiwan has the national contact tracing platform TRACE developed by CDC in 2018 [9,10]. In addition, Taiwan CECC also set up a smartphone-based real-time locating system to track contacts’ phone signals and alert local authorities if anyone left their designated location or switched off their phone [9]. All individuals determined to have been in contact with an infected person must quarantine at home for 14 days. This data has also assisted officials in ensuring that individuals remain quarantined for the required period. Taiwan’s experience with past epidemics (i.e., SARS in 2003 and H1N1 in 2009) has influenced Taiwan CDC to continuously improve and adjust strategies for pandemic response and control before vaccination program [11].

The next wave during May–July 2021 was the major outbreak that took place in Taiwan after more than a year of COVID-19 (450–530 domestic cases reported at the end of May). Taiwanese production of medical masks, a high percentage of mask-wearing, alcohol sanitizer, other medical supplies, and establishment of a QR-code registration system at stores and other establishments for contact tracing played the most important role to eliminate the outbreak during a few months. In addition, another important non-pharmaceutical intervention was fast and strict restrictions on mass gathering: five people indoors and ten people outdoors [12]. It should be noted that the strict gathering restriction was in action from May 19 to July 27. After July 27, the allowed number of gathering people has been increased to 50 indoors, 100 outdoors [13]. By July 2021, daily numbers of domestic cases returned to the single digits for the first time. August 25 saw the first day with zero cases since the start of the outbreak. From August 24, limit on the number of people in gatherings raised to 80 people indoors and 300 people outdoors [14], and only after October 5, some entertainment venues had been opened [15]. Pharmaceutical intervention – vaccination – has been only begun in 2021. On March 22, 2021, the first vaccination program using AstraZeneca vaccine was started in Taiwan [16]. On July 27, 2021, people with two doses were 1.2% and people with one dose were 28% of the whole population of Taiwan [17].

During the wave in January–March 2022, only non-pharmaceutical interventions were not enough to contain the outbreak. New variants Delta and Omicron have much higher infection rates than previous variants. For outbreak control, vaccination is also necessary together with other non-pharmaceutical interventions. However, vaccination has smaller protection against Omicron than Delta [18]. In Taiwan, on January 13, 2022, already 71.55% was vaccinated with the second dose and only 3.46% with booster, whereas on March 7, 2022, 77.22% was with the second dose and 44.84% with booster [17,19]. Recent studies investigated that vaccination after the second dose loses its efficiency after 20 weeks but a booster dose can again increase the efficiency by more than 60% [18,20–23]. This is why Taiwan started to gradually relax various gathering restrictions from March 2022, since pharmaceutical interventions (vaccination) were implemented at high levels [24]. However, Taiwan’s success so far is no guarantee that it can totally control COVID-19, and the government continues to urge people to remain vigilant, continue wearing masks, and do vaccination as the pandemic continues.

Considering the above-mentioned interventions as well as the flow of travelers between Wuhan and Taiwan from December 2019 to January 2020, we simulated different scenarios of COVID-19 waves in Taiwan from January 2020 to the beginning of March 2022. The model results demonstrate the effectiveness of the rapid implementation of various control measures for reducing local transmission during a pandemic before a universal pharmaceutical intervention (from 2020 to March 21, 2021) and together with pharmaceutical intervention (from March 22, 2021 to the beginning of March, 2022).

## 2. Materials and methods

### 2.1. Model

The susceptible-exposed-infectious-removed (SEIR) model is frequently used for modeling disease dynamics [25–27]. With the standard SEIR model, individuals are placed in one of four stages: susceptible (S), exposed (E), infected (I), and removed (or recovered) (R). However, it is now well-reported that pre-symptomatic and asymptomatic cases are crucial in the spread of COVID-19, and several modified versions of the SEIR model have been proposed [28–31]. Modifications include the addition of asymptomatic cases [28,31], a special category of individuals who are quarantined or hospitalized [30,31], and the consideration of contact tracing and mask-wearing [29–31]. In the current study, we propose an epidemiological compartmental model that accounts for all of the above-mentioned modifications. We hypothesize that this modified model can more precisely describe the dynamic transmission of COVID-19 compared to a simple SEIR model. We used the modified model to describe the dynamics of the spread of COVID-19 in Taiwan during the 2020–2022 years and demonstrate the impact of various interventions especially such as mask-wearing and contact tracing. In addition, we developed a formula for the basic reproduction number (*R*_0_) for our model, which describes the dependence of *R*_0_ on all model parameters. To define *R*_0_, we used the next-generation matrix (NGM) approach proposed by Diekmann et al. [32,33]. It should be mentioned that in the current study we use fixed averaged values for the parameters and don’t use any distributions for the model parameters.

#### 2.1.1. Model including asymptomatic and pre-symptomatic cases

To ensure our model accurately reflects the real-world scenario, we added asymptomatic cases and pre-symptomatic cases to the modified SEIR model, based on previous models [28,31]. This initial model includes six stages: susceptible (S), asymptomatic (A), exposed or pre-symptomatic (E), infected or symptomatic (I), symptomatic under quarantine or hospitalized (Q), and recovered (R). The following system of equations describes the model:

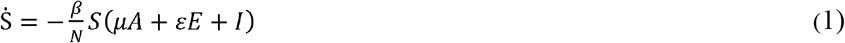

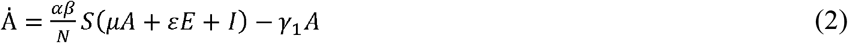

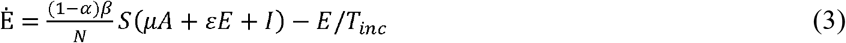

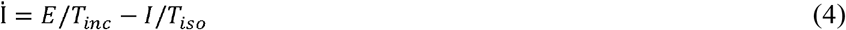

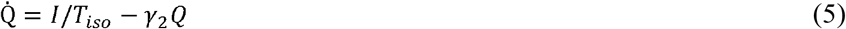

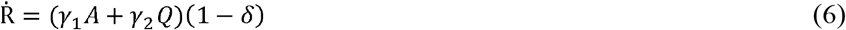

Eq (1) describes the dynamics of reducing the number of uninfected (susceptible) people *S*. The transmission rate of the COVID-19 from infected cases denoted *β*, is one of the most important parameters. Asymptomatic (*A*) and pre-symptomatic (*E*) cases have smaller transmission abilities than infected persons with symptoms (*I*). Therefore, we have the relative infectivity of asymptomatic individuals denoted *μ*, and the ratio of pre-symptomatic transmission (the probability of transmission before symptoms appear) denoted *ε*.

Eq (2) describes the dynamics of asymptomatic cases *A*. Each person in the *S* state can be converted to either the *A* state with probability *αβ* (where *β* is the transmission rate and *α* is the probability of being asymptomatic after infection). An asymptomatic individual infects others with probability *αβ* and recovers with probability *γ*_1_ = 1/*T*_*a,inf*_,, where *T*_*a,inf*_ is the duration of infection for asymptomatic individuals.

Eq (3) describes the dynamics of exposed individuals *E* who are infected but have not yet developed symptoms and, thus, are pre-symptomatic. The probability that an individual is susceptible and will later become pre-symptomatic is (1 − *α*)*β*. A person in the *E* state will move to the *I* state with probability 1/ *T*_*inc*_, where *T*_*inc*_ is the incubation period.

Eq (4) describes the dynamics of symptomatic infected people *I* who are not yet hospitalized or quarantined and can infect healthy people *S*. People in the *I* state can be isolated with a delay of *T*_*iso*_ between the onset of symptoms and isolation. *T*_*iso*_ is called isolation time. Therefore, an individual from the *I* state will move to the *Q* state with probability 1/ *T*_*iso*_.

Eq (5) describes the dynamics of symptomatic infected people under home quarantine or hospitalization (*Q*). People in the *Q* state will recover (move to the *R* state) or die with probability *γ*_2_ = 1/(*T*_*r*_ − *T*_*iso*_), where *T*_*r*_ represents the interval between the onset of symptoms and recovery or death. The difference between *T*_*r*_ and *T*_*iso*_ is the interval between hospitalization and recovery or death.

Eq (6) describes the dynamics of people who have recovered and are no longer contagious (*R*). Since this stage represents only recovered people, all people in the *R* state will be alive with probability (1− *δ*), where *δ* is the death rate.

#### 2.1.2. Model including contact tracing

On January 15, 2020, the Taiwan Centers for Disease Control (Taiwan CDC) made COVID-19 a notifiable disease [34]. Therefore, we can assume that contact tracing began in Taiwan on this date. Based on work by Nuzzo et al. [30], to account for this intervention in the model, we introduce the parameter *tr* which represents the proportion of traced contacts. A new state accounts for people who were in contact with an infected person and isolated as a result of contact tracing (therefore, can no longer infect others). Considering, there are asymptomatic and pre-symptomatic cases within the *C* state, we divide it into two parts:

- *C*_*s*_ comprises pre-symptomatic people from the *E* state who are self-isolating due to contact tracing;
- *C*_*a*_ comprises asymptomatic people from the *A* state who are self-isolating due to contact tracing.

Both pre-symptomatic and asymptomatic cases may be under self-isolation if they were in contact with a symptomatic infected person. In other words, if “parents” of new cases in the *E* or *A* states (from whom they were infected) are symptomatic then contact tracing can be done for these new cases. For pre-symptomatic individuals in the *E* state, contact tracing works but with some delay. We suppose that after the onset of symptoms in one person, their traced contacts will be under self-quarantine with a delay *T*_*iso*_. We assume this delay because of the time required to receive COVID-19 test results. Therefore, a person in the *E* state can move to the *C*_s_ state with probability 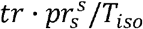, where 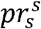 is the probability that the “parent” of the new case is symptomatic. A person in the *A* state can move to the *C*_*a*_ state with probability 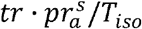, where 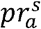 is the ratio of new asymptomatic cases produced from symptomatic.

We suppose that after self-isolation, asymptomatic infected people will not be infected again, so individuals in the *C*_*a*_ state move to the *R* state after a home quarantine period *T*_*q*_ with probability *γ*_3_ = 1/*T*_*q*_ A person from the *C*_s_ state can move to the *Q* state after the development of symptoms (since they were traced in the pre-symptomatic period). The time delay *T*_*iso*_ must also be considered. Therefore, the probability that a person will move from the *T*_*s*_ state to the *Q* state is *γ*_4_ = 1/(*T*_*inc*_ − *T*_*iso*_). When contact tracing measures are included, our model takes the following form:

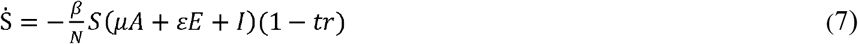

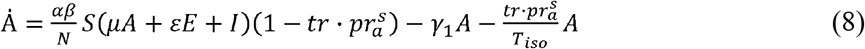

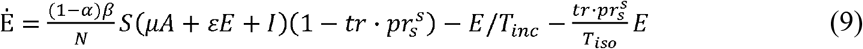

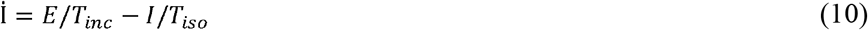

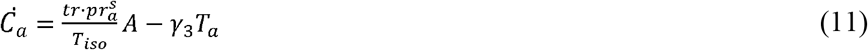

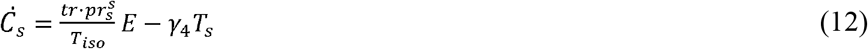

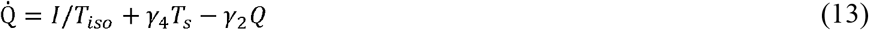

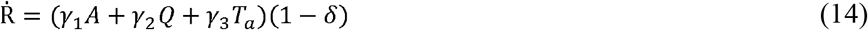

#### 2.1.3. Model including mask-wearing

Mask-wearing is one of the main control measures to combat the spread of COVID-19 transmission [35–37]. To account for mask-wearing in our model, we suppose that masks are worn by a certain percentage *p* of the total population *N* and are not worn by a 1 − *p* percentage of the total population *N*. We also suppose that a mask can reduce both the spread of germs and droplets from an infected person and protect a healthy person from these germs with different effectiveness [38,39]. Mask efficiency *e* depends on its type (medical or homemade), the number of layers, and the material [40–44]. For example, the effectiveness of homemade masks is estimated to range from 2% to 38% whereas the efficiency of surgical masks can vary from 40% to 90% [34,43,45,46]. In our model, *r* = 1 − *e* is used to describe the reduction in the probability of contagion from one person wearing a mask in an *S*–*A, S*–*E*, or *S*–*I* contact [29].

Based on the previous studies [29,31], the subscript *n* denotes the people who don’t wear a mask, and the subscript *m* denotes the people who wear a mask. Therefore, the number of equations in the system is doubled. Denoted by *X* = *μA*_*n*_ +*rμA*_*m*_ + *εE*_*n*_ + *rεE*_*m*_ + *I*_*n*_ + *rI*_*m*_, the model that includes mask-wearing has the following form:

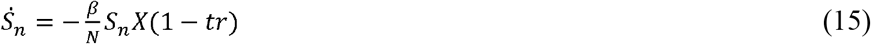

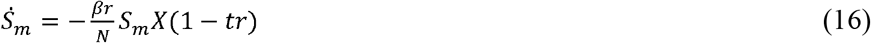

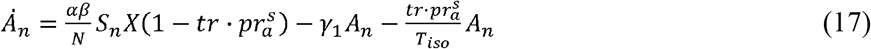

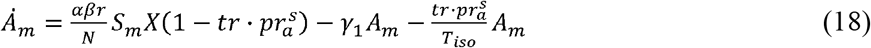

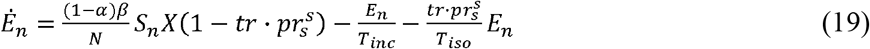

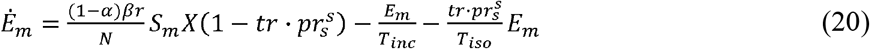

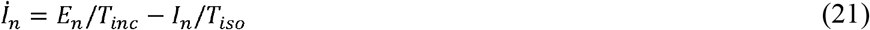

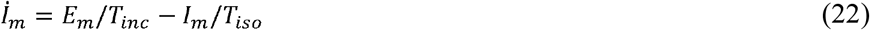

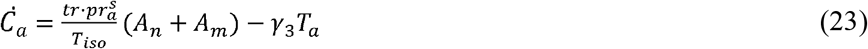

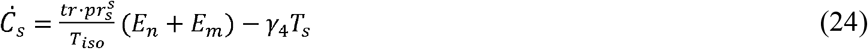

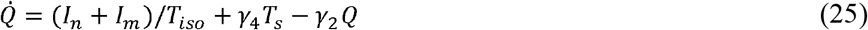

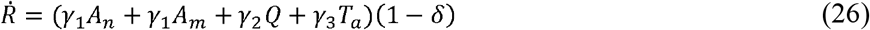

The obtained system of equations (15)–(26) is used to simulate the COVID-19 wave in Taiwan during 2021–2022 years. To simulate the beginning of the COVID-19 in Taiwan in January–March of 2020 year, it is necessary to include in the model travelers from Wuhan.

#### 2.1.4. Model including travelers

Since the majority of cases in Taiwan in January 2020 were imported from Wuhan, we model the number of infected people in Wuhan using the SEIR model, taking into account foreign and local (within China) travelers [25]. We use this separate model for travelers to make our final model more realistic.

We denote the estimated number of non-infected and infected passengers from Wuhan to Taiwan as *P*_*WT*_ and 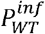, respectively (for a detailed explanation of the Wuhan model and the derivation of formulas see Supplementary Materials).

Because there is no data about how many travelers were asymptomatic or latent, we added 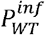 to the *A, E*, and *I* states. We also split non-infected travelers *P*_*WT*_ into two groups: those who wear masks (with probability *p*) and those who don’t wear masks (with probability *1* − *p*). Adding *P*_*WT*_ and 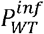 to the Eqs (15)–(22), we obtain the model describing the dynamics of COVID-19 propagation in Taiwan during January–March of 2020 year:

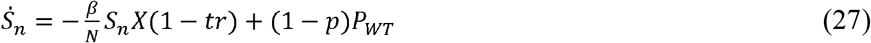

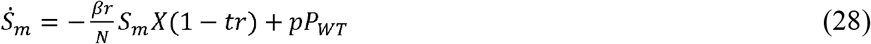

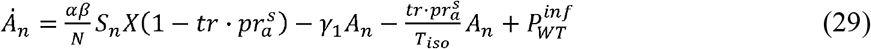

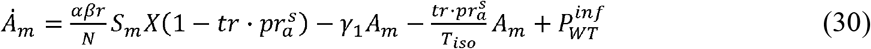

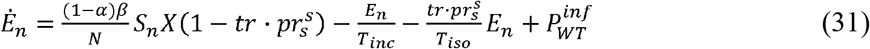

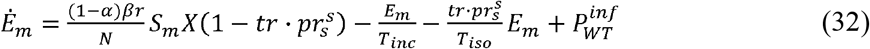

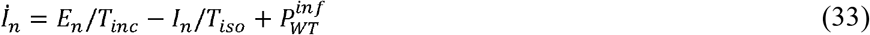

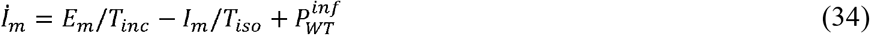

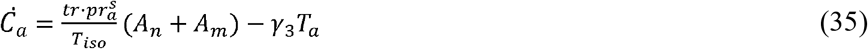

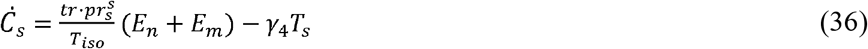

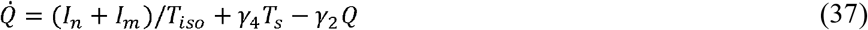

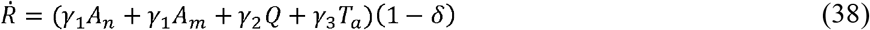

### 2.2. Simulation parameters

In the current section, the values of all model parameters are described for the different COVID-19 waves in Taiwan. Table 1 displays the default values of all parameters used in the model, and Table 2 displays the all non-pharmaceutical interventions.

**Table 1.**
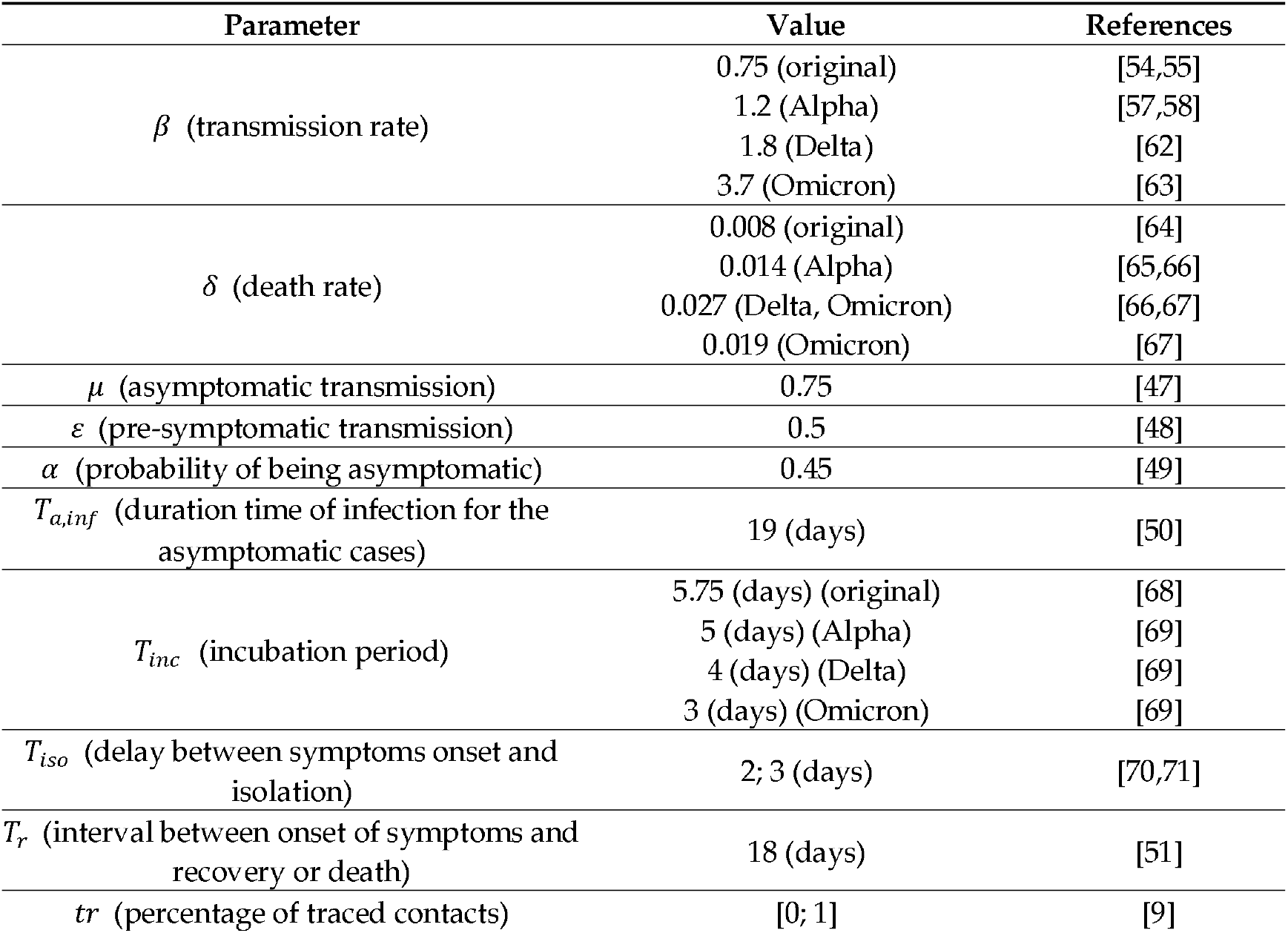
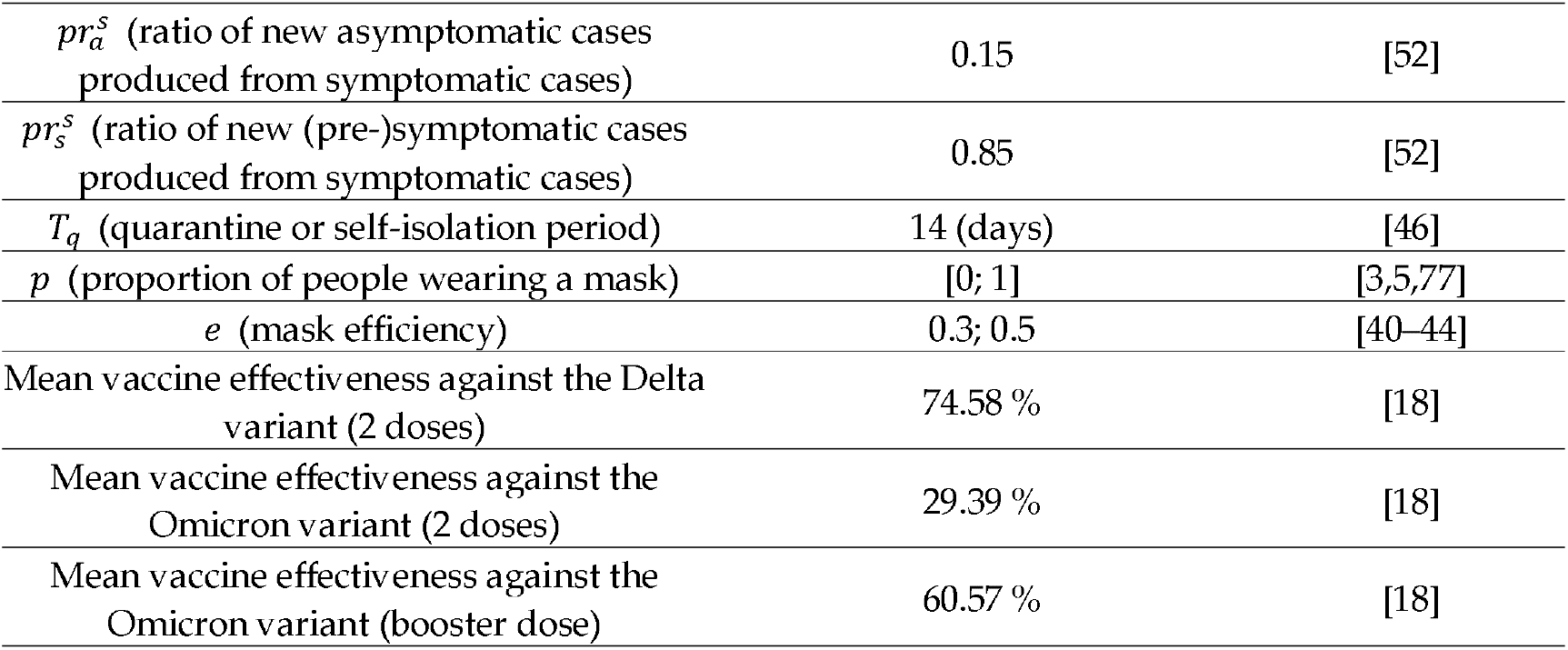
The parameters used in the models.

| Parameter | Value | References |
| --- | --- | --- |
| $\beta$ (transmission rate) | 0.75 (original) | [54,55] |
|  | 1.2 (Alpha) | [57,58] |
|  | 1.8 (Delta) | [62] |
|  | 3.7 (Omicron) | [63] |
| $\delta$ (death rate) | 0.008 (original) | [64] |
|  | 0.014 (Alpha) | [65,66] |
|  | 0.027 (Delta, Omicron) | [66,67] |
|  | 0.019 (Omicron) | [67] |
| $\mu$ (asymptomatic transmission) | 0.75 | [47] |
| $\varepsilon$ (pre-symptomatic transmission) | 0.5 | [48] |
| $\alpha$ (probability of being asymptomatic) | 0.45 | [49] |
| $T_{a,inf}$ (duration time of infection for the asymptomatic cases) | 19 (days) | [50] |
| $T_{inc}$ (incubation period) | 5.75 (days) (original) | [68] |
|  | 5 (days) (Alpha) | [69] |
|  | 4 (days) (Delta) | [69] |
|  | 3 (days) (Omicron) | [69] |
| $T_{iso}$ (delay between symptoms onset and isolation) | 2; 3 (days) | [70,71] |
| $T_r$ (interval between onset of symptoms and recovery or death) | 18 (days) | [51] |
| $tr$ (percentage of traced contacts) | [0; 1] | [9] |
| $pr_a^s$ (ratio of new asymptomatic cases produced from symptomatic cases) | 0.15 | [52] |
| $pr_s^s$ (ratio of new (pre-)symptomatic cases produced from symptomatic cases) | 0.85 | [52] |
| $T_q$ (quarantine or self-isolation period) | 14 (days) | [46] |
| $p$ (proportion of people wearing a mask) | [0; 1] | [3,5,77] |
| $e$ (mask efficiency) | 0.3; 0.5 | [40–44] |
| Mean vaccine effectiveness against the Delta variant (2 doses) | 74.58 % | [18] |
| Mean vaccine effectiveness against the Omicron variant (2 doses) | 29.39 % | [18] |
| Mean vaccine effectiveness against the Omicron variant (booster dose) | 60.57 % | [18] |

**Table 2.** Starting dates of interventions and stages.

| Interventions | Start dates | References |
| --- | --- | --- |
| **January–March 2020** |  |  |
| 30% of people wear masks | December 25, 2019 | [3,5,77] |
| Contact tracing program | January 15, 2020 | [34] |
| 50% of people wear masks | January 22, 2020 | [3,5,77] |
| Start of Wuhan lockdown | January 23, 2020 | [82] |
| 70% of people wear masks | January 31, 2020 | [3,5,77] |
| 90% of people wear masks | February 6, 2020 | [3,5,77] |
| **May–July 2021** |  |  |
| Strong gathering restrictions in Taipei City and New Taipei City | May 15, 2021 | [72] |
| Reducing of isolation time to 2 days | May 15, 2021 | [83] |
| 90% of people wear masks | May 15, 2021 | [72] |
| 70% of contacts are traced | May 15, 2021 | [72,83] |
| Nationwide strong gathering restrictions | May 19, 2021 | [12] |
| 99% of people wear masks | May 19, 2021 | [12] |
| 90% of contacts are traced | May 19, 2021 | [12] |
| Nationwide moderate gathering restrictions | July 27, 2021 | [13] |

#### 2.2.1. Fixed parameter values for all waves

The relative infectiousness of asymptomatic individuals *μ* is difficult to investigate although the Centers for Disease Control and Prevention (CDC) estimated that *μ* = 0.75 [47]. Based on work by He et al. [48], the ratio of pre-symptomatic transmission *ε* = 0.45. Estimation of the proportion of asymptomatic cases is difficult, however, based on the results of a recent study [49], the probability of being asymptomatic after infection is assumed *α* = 0.45. The duration of infection for asymptomatic individuals *T*_*a,inf*_ is around 19 days [50]. According to the clinical characteristics of COVID-19 [51], *T*_*r*_ = 18 days. He et al. estimated the ratio of new asymptomatic cases produced from symptomatic 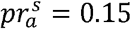 [52]. Thus, we set 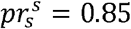. The home quarantine period *T*_*q*_ = 14 days.

In this paper, we simply assume the main mask efficiency *e* = 0.5 since, in Taiwan, the majority of people wear medical masks since their access is guaranteed and facilitated by the government [2,53]. Further, medical masks were available at a fixed price from the government beginning in early March 2020 in Taiwan. To investigate how mask effectiveness can affect the results, we also consider lower mask efficiency *e* = 0.3 (but only during 2020 year, since during 2020, the national mask production was being adjusted to provide all residents with medical masks [3,8], and people could wear non-medical masks).

#### 2.2.2. Different parameter values for different waves

Value of the transmission rate *β* depends on the COVID-19 variant. For the original COVID-19, *β* estimation varies from 0.5 to 1 [54,55], so we took 0.75. The Alpha variant (B.1.1.7) of the COVID-19 (that was during spring 2021 outbreak in Taiwan [56]) is estimated to be 40–80% more transmissible than the original COVID-19 version [57–61]. We assumed that the transmission rate for the Alpha variant is 60% higher than original COVID-19 and *β* = 1.2 [57,58]. The Delta variant (B.1.617.2) estimation demonstrated that it can be 50% more transmissible than the Alpha variant and *β* = 1.8 [62]. For the Omicron variant (B.1.1.529), the transmission rate estimated to be 105% higher than the Delta variant and *β* = 3.7 [63].

The death rate *δ* also depends on the COVID-19 variant. For the original COVID-19, *δ* is estimated between 0.002 and 0.015 [64] and we assumed *δ* = 0.008. For Alpha variant, we consider *δ* = 0.014 [65,66]. For Delta variant, we take *δ* = 0.027 [66,67], and for Omicron variant, *δ* = 0.019 [67].

The incubation period *T*_*inc*_ also depends on the COVID-19 variant. *T*_*inc*_ decreases for each new COVID-19 variant. According to recent studies, for the original COVID-19 variant *T*_*inc*_ = 5.75 days [68]; for the Alpha variant *T*_*inc*_ = 5, for the Delta variant *T*_*inc*_ = 4, for the Omicron variant *T*_*inc*_ = 3 [69].

From the previous studies, the mean value of the isolation time *T*_*iso*_ varies between three and four days [70,71]. During the spring outbreak in 2021, Taiwan started to set up outdoors rapid testing posts [72] which is able to reduce the isolation time. We use *T*_*iso*_ = 3 days for the wave in January–March 2020 and in the beginning of the spring wave. *T*_*iso*_ = 2 is used during the 2021–2022 years.

Probability of mask-wearing *p* and contact tracing *tr* is varied from 0% to almost 100% for different time periods. A study by Jian et al. [9] found that with tracing apps and automatic text messages, as in Taiwan, the ratio of detected cases via contact tracing was 61.5% (during 2020 year). Another study shows [73], during 2020 year around 81% of Taiwanese citizens wore masks outdoors and around 99% of people wore masks indoors or while taken mass transportation.

#### 2.2.3. Mass gathering restrictions efficiency

Mass gathering restriction is one of the most powerful non-pharmaceutical interventions that may relatively fast decrease the disease transmission. As WHO says in its guidance about planning of mass gatherings [74], the high density and mobility of attendees associated with mass gatherings can entail a higher risk of transmission of COVID-19 and a potential destruction of the health system’s response capacities if large numbers of people are infected.

Mass gathering restrictions can give different strength: light restrictions, for example, gatherings of over 500 people [24,75], moderate restrictions as 50 people indoors and 100 people outdoors [13], or very strict restrictions like five people indoors and ten people outdoors [12,72]. Since mass gathering restrictions can reduce the COVID-19 transmission, therefore, the efficiency of gathering restrictions decreases the value of the transmission rate *β*. In the current paper, we considered that the strict mass gathering restrictions have 46% efficiency which was estimated based on the authors previous work [76]. Other types of the gathering restrictions were assumed with the following efficiencies: moderate strength gathering restrictions have 20% efficiency, light strength gathering restrictions have 15% efficiency.

#### 2.2.4. Vaccination efficiency

Recent studies demonstrated that two doses are more effective than one dose but vaccine efficiency is reduced during time [18,20–22]. Moreover, vaccines have much smaller resistance to the Omicron variant than to Alpha or Delta variants. However, the booster dose is capable to increase the vaccine effectiveness which can strongly decrease after 15–20 weeks interval after the second dose vaccination [18]. Efficiency of vaccine also depends on vaccine type [18,20–22]. Based on the detailed investigation of different vaccines effectiveness against the Delta and Omicron variants [18], the average value of vaccine efficiency was calculated. The vaccination efficiency by two doses has 74.58% against the Delta. Against the Omicron, two doses vaccination has 29.39% efficiency whereas booster dose has 60.57%.

Vaccination for the spring outbreak in 2021 year (Alpha variant) was not taken into account since fewer than 1% of Taiwan’s residents had been vaccinated [17,19]. The low vaccination rate was caused by supply issues with purchased overseas vaccines and the calming of the Taiwanese population due to low case numbers during the 2020 year.

## 3. Results

### 3.1. January–March 2020 (original COVID-19 variant)

We modeled the wave of COVID-19 transmission in Taiwan during January–March 2020. We supposed that the percentage of the mask-wearing population in Taiwan increased as the government facilitated universal access to masks. While it is difficult to estimate the percentage of the population who wear masks, our assumptions were based on measures taken by the Taiwanese government and Taiwan CDC and the habitual mask-wearing that is part of the culture in some Asian countries, including Taiwan [3]. Taiwanese people are accustomed to wearing masks when they are ill [53], and there is recent evidence of widespread adherence to mask-wearing. According to a survey among fifth-to twelfth-grade students in Taiwan on the impact of COVID-19 [77], around 63% wear a mask all the time and 25.5% wear a mask in public places, which means around 90% of adolescents in Taiwan wear a mask in public places.

In our model, we included and excluded various interventions (listed in Table 2 along with implementation dates). Figures 2–3 show the daily numbers of infected symptomatic individuals and asymptomatic individuals. Running the model starts from December 25, 2019, with the following initial values:,,.

**Figure 2.**
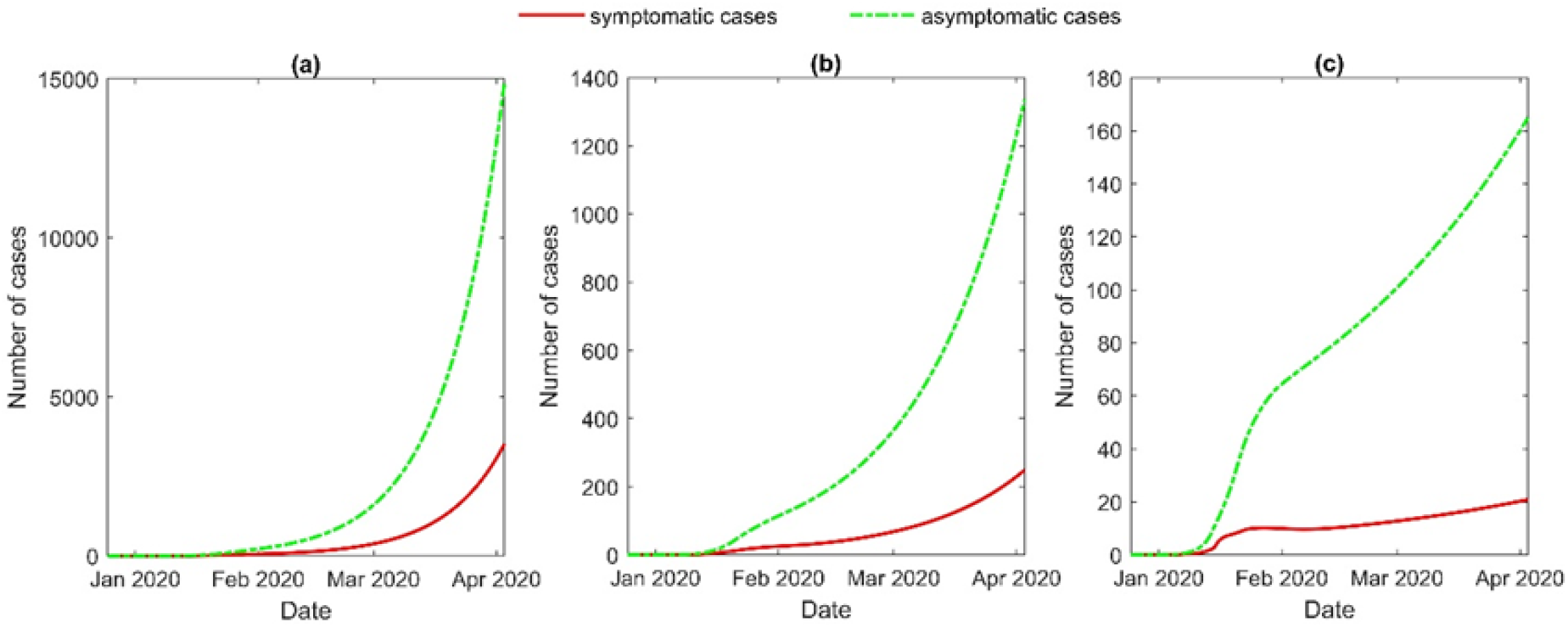
Results of 30% mask efficiency and different percentages of traced contacts: **(a)** 40%; **(b)** 60%; **(c)** 80%.

We varied the percentage of traced contacts and the effectiveness of masks to obtain the simulation curve of symptomatic cases that is close to official COVID-19 data for the January–March period and to show the influence of these interventions. Figures 2a and 3a show the results when 40% of contacts are traced. Using to represent low effectiveness of masks, the number of cases increases until the beginning of April, with the number of symptomatic cases reaching around 4,000 (Figure 2a). Using to represent the effectiveness of medical masks, the curve peaks around the beginning of February (Figure 3a), after which there is a slow decrease. The behavior of curves with efficiency and other values for contact tracing, or (Figure 2b–c) is approximately the same as in Figure 2a, but the maximum number of cases is smaller by tens (Figure 2b) and hundreds of times (Figure 2c). For and (Figure 3b) or (Figure 3c), the maximum number of cases is around ten and five, respectively. In addition, after peaking, the curves in Figure 3b–c move toward zero faster than the curve in Figure 3a.

**Figure 3.**
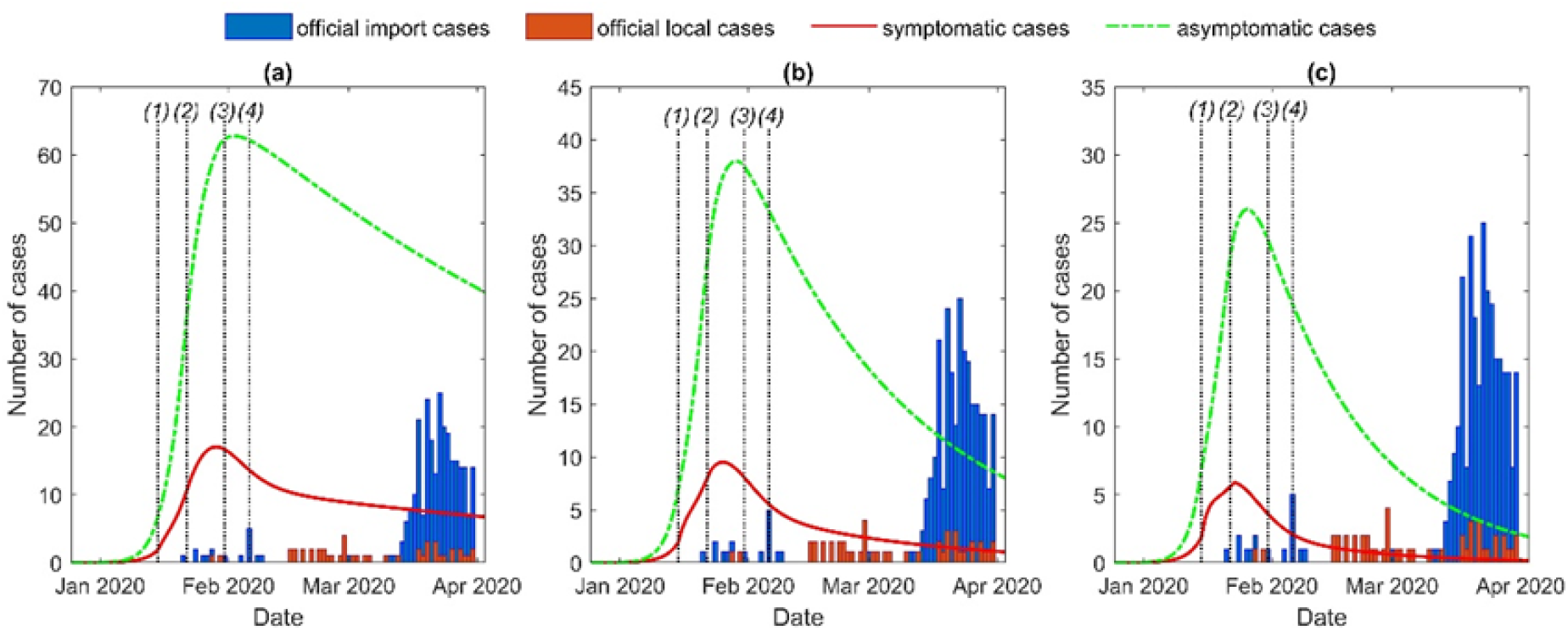
Comparison of the number of officially reported cases with the simulated results. In the model, 50% mask efficiency and different percentages of traced contacts are considered: **(a)** 40%; **(b)** 60%; **(c)** 80%. The main interventions (dates are listed in Table 2): *(1)* start of contact tracing; *(2)* 50% of people wearing masks and start of Wuhan lockdown; *(3)* 70% of people wearing masks; *(4)* 90% of people wearing masks.

In Figure 3b–c, the results of our model are very close to the official data before the wave of imported cases from Europe and the United States at the end of March 2020. Moreover, our model can estimate the approximate number of asymptomatic cases during January–March 2020 wave in Taiwan. Using data from Figure 3a, the maximum number of asymptomatic cases was around 60 at the beginning of February 2020.

In addition, we estimated by using the NGM method (see Supplementary Materials). Figure 4 represents the dependency of on contact tracing and mask-wearing probability.

**Figure 4.**
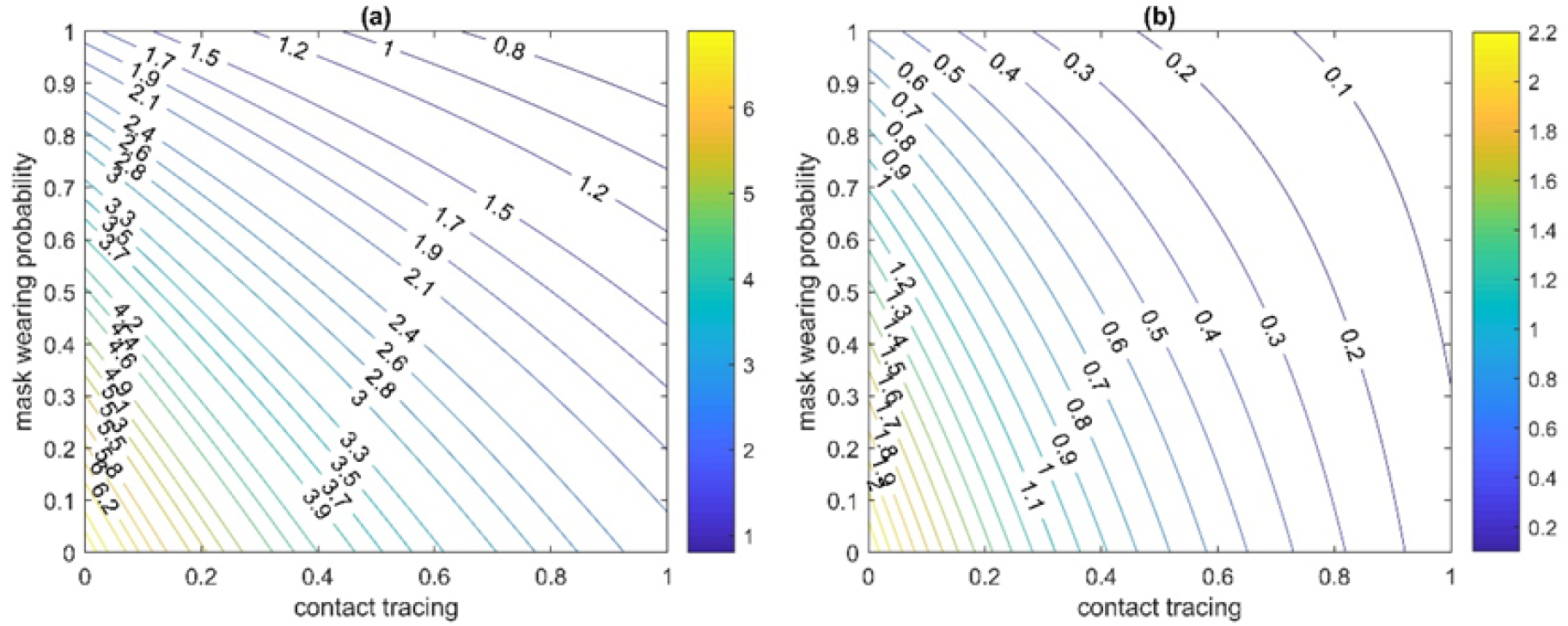
Estimation of R_0_, dependent on mask-wearing probability and contact tracing (using the next-generation matrix approach) with consideration of asymptomatic cases **(a)** and with only consideration of symptomatic cases **(b)**.

In the model described in Eqs (27)–(38), many parameters can affect. However, in this paper, we focused on the percentage of traced contacts and the percentage of people wearing masks. Since our model include asymptomatic cases, was estimated with and without asymptomatic cases (see Supplementary Materials). with consideration both asymptomatic and symptomatic cases (Figure 4a) can be much higher than with only consideration of symptomatic cases (Figure 4b). Figure 4a represents that to reach below one, the percentage of both traced contacts and people wearing masks must be relatively high, for example, for and,. Figure 4b shows that can be less than one, and, therefore, the outbreak can be controlled when both contact tracing and mask-wearing probability are greater or equal to 30% (for and,. However, such a big difference between results with and without asymptomatic cases demonstrates how relatively easy the outbreak would be controlled without asymptomatic cases. In the following sections, the full NGM approach with asymptomatic cases consideration will be used for estimations of.

### 3.2. May–July 2021 (Alpha variant)

To model the spring 2021 outbreak of COVID-19 in Taiwan during May–July 2021, we supposed that during April and the first half of May, the proportion of the mask-wearing population was sufficiently high and equal to 80% [72], whereas contact tracing was assumed to be equal to 50%, since small number of local cases during whole 2020 year had some relaxation effect.

When the number of cases started to increase very fast, the government introduced very quickly strict interventions such as mandatory mask wearing at all times when outside, shutting cinemas and entertainment spots, and gathering restrictions to five indoors and ten outdoors [12,75]. All interventions with dates and their impact on the model parameters are described in Table 2. Simulation of the outbreak in 2021 year starts from April 1, 2021 with the following initial conditions:,,,,.

Figure 5a demonstrates comparison of official data with simulation results. The obtaining results proves that the fast response of the government with strict interventions is capable to relatively quickly controlling the outbreak. At the peak of the outbreak, the mask-wearing probability was 99% and contact tracing was 90%. Figure 5c shows that estimated is very fast dropped below one that allows to reduce number of new cases also relatively fast. Even after some relaxation on gathering restrictions at the end of July (50 people indoors and 100 outdoors) [13], is still less than one that can guarantee outbreak control.

**Figure 5.**
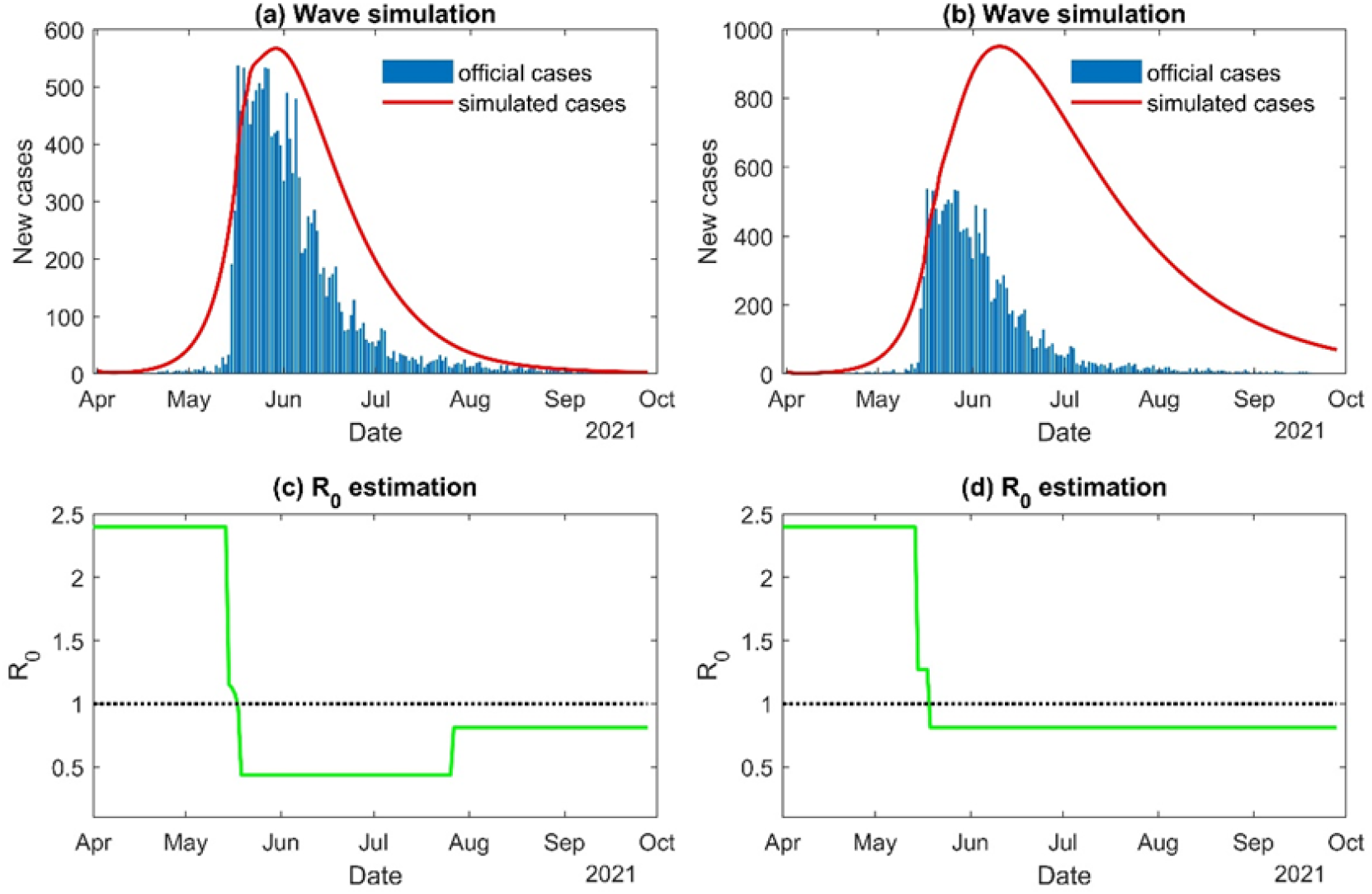
Simulation wave with R_0_ estimation when all interventions are used **(a)** and **(c)** and when only contact tracing and mask wearing are used **(b)** and **(d)**. Interventions and their dates are listed in Table 2.

Figures 5b and 5d represent the scenario when only probability of mask wearing and contact tracing is increased without mass gathering restrictions. We can see that number of cases is greater almost twice in comparison with official data, and the decrease of number of cases happens much slower than official numbers (Figure 5b). In this scenario, it is still possible to reach outbreak control, however, not so fast and with a much higher number of cases. In Figure 5d, it can be seen that rapid increase of contact tracing (*tr* = 0.9) and mask-wearing (*p* = 0.99) is able to reduce the *R*_0_ value below one.

### 3.3. July–December 2021 (Delta variant)

During the second half of 2021, the domestic number of cases was close or equal to zero, despite the fact that the Delta variant had been spread globally in this period. Taiwan continued to adhere to pandemic prevention guidance in order to save the outbreak control. Since Taiwan has only started the vaccination program on March 2021, strong gathering restrictions were almost until the end of 2021 [14,15], mask-wearing and contact tracing also performed in a high level.

We estimate the value of the basic reproduction number *R*_0_ by using the NGM method to demonstrate how Taiwan was able to keep the local number of cases at zero level.

Figure 6 represents the dependence of the *R*_0_ on the mask-wearing and contact tracing probabilities but without consideration of any mass gathering restrictions. In Figure 6, there are the different proportions of vaccinated people by two doses, since from July to November 2021, the proportion of the fully vaccinated population had increased from 1.2% to 50% [17,19]. From Figure 6a–b, it can be seen that the outbreak control (*R*_0_ < 1) is impossible with 0–25% of vaccinated people, even with perfect quality of mask-wearing and contact tracing. With a higher proportion of vaccinated people, 50–75% (Figure 6c–d), the outbreak can be controlled, however, only with the extremely high-level implementation of mask-wearing and contact tracing.

**Figure 6.**
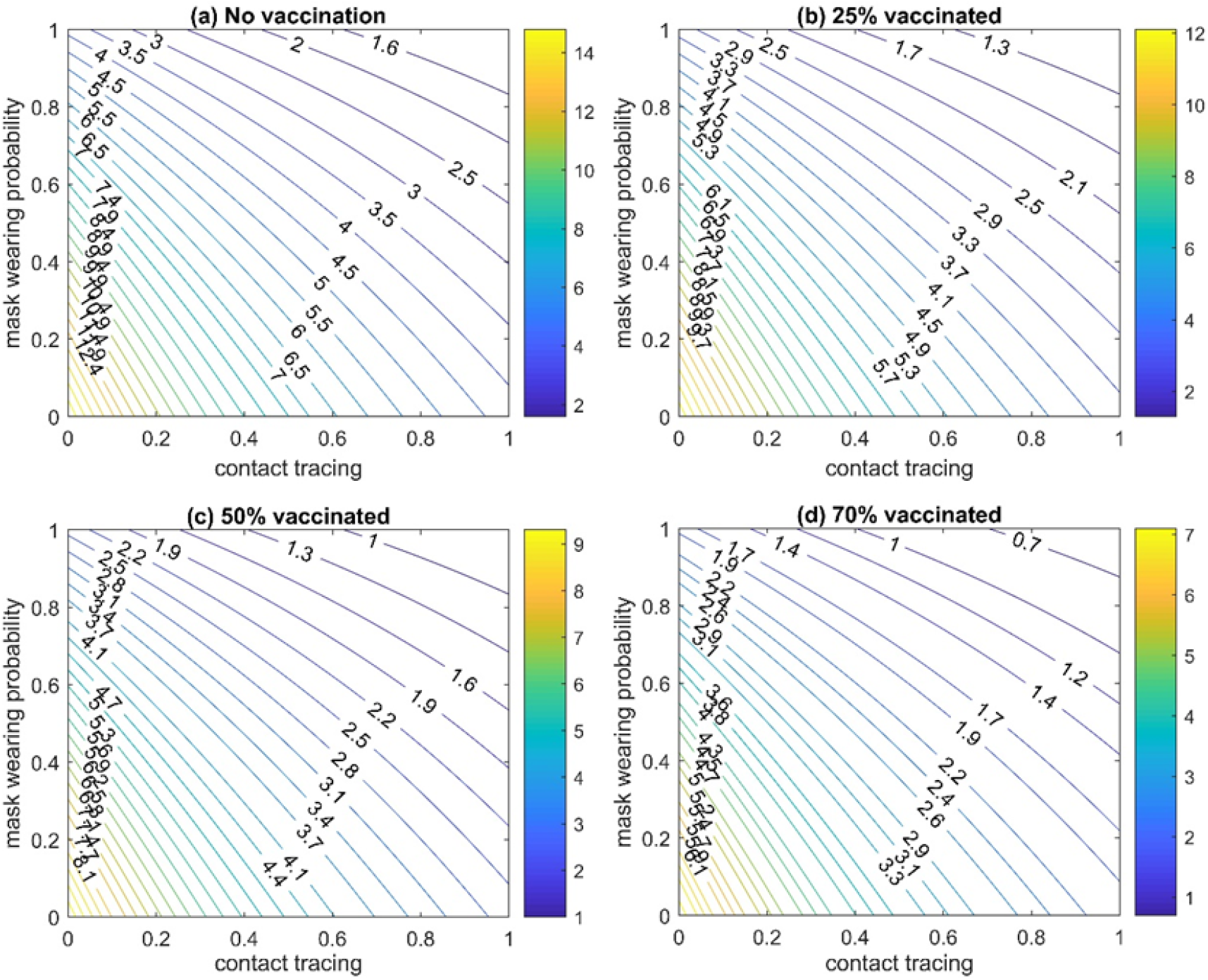
The basic reproduction number (R_0_) estimation for Delta variant without mass gathering restriction and with different proportion of vaccinated people by two doses: **(a)** 0%; **(b)** 25%; **(c)** 50%; **(d)** 70%.

Figure 7 demonstrates the estimation also for different proportions of vaccinated population but with consideration of mass gathering restrictions along with mask-wearing and contact tracing. In Figure 7, it can be seen that can be less than one (the outbreak is under control) for all different considered percentages of vaccinated people (from 0% to 70%). For 0–25% of vaccinated people (Figure 7a–b), the control of outbreak is possible only when mask-wearing and contact tracing are realized more than 80%. Figure 7c shows that already with 50% of the vaccinated population, an execution of mask-wearing and contact tracing on the medium level is enough to reach smaller than one. In Figure 7d, the outbreak is controlled even without mask-wearing and contact tracing. Note, Figures 6–7 present the results for the Delta variant. As it will be shown further, for the Omicron variant outbreak control, higher proportions of the vaccinated population together with the booster vaccine program are needed.

**Figure 7.**
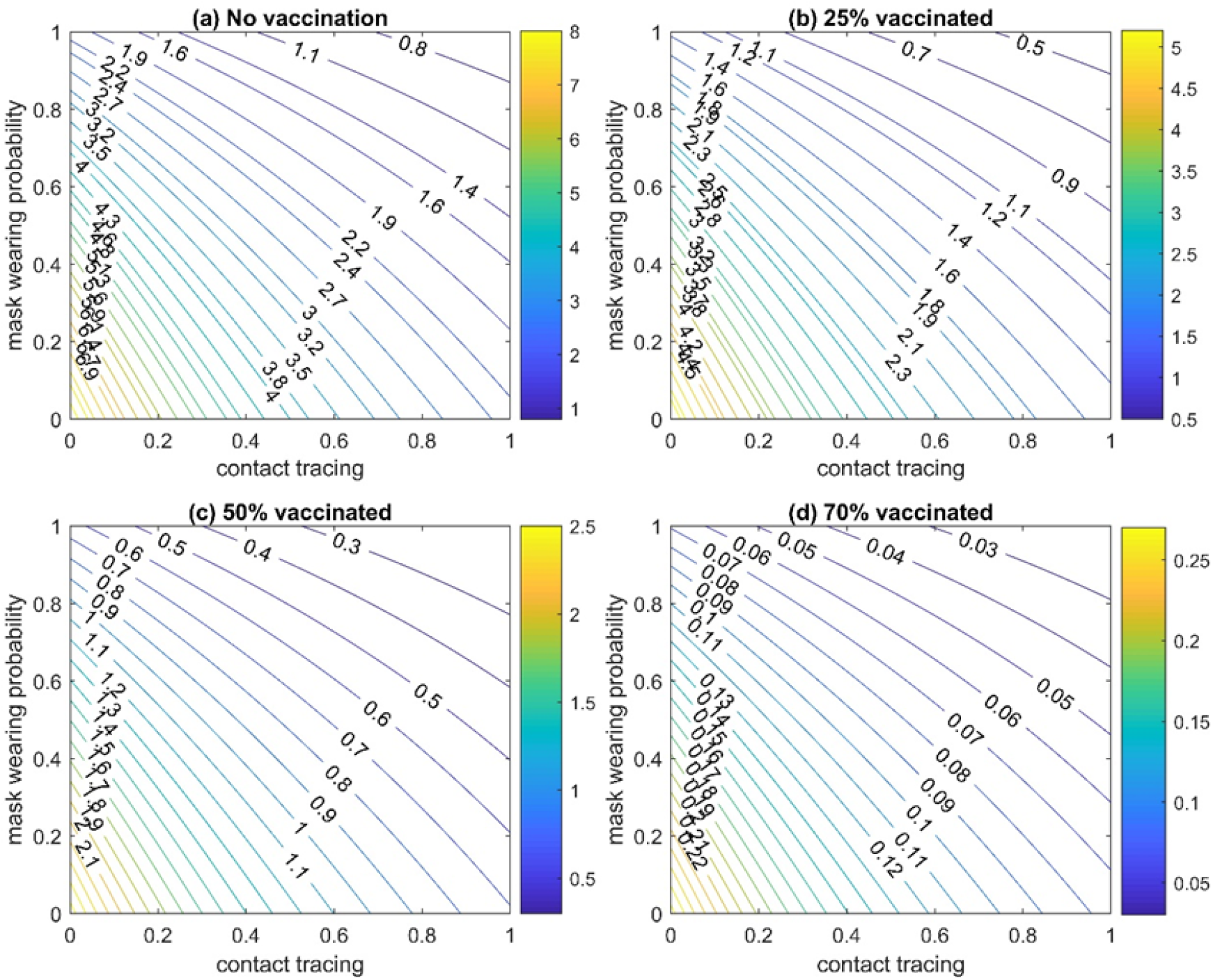
The basic reproduction number (R_0_) estimation for Delta variant with strong mass gathering restriction and with different proportion of vaccinated people by two doses: **(a)** 0%; **(b)** 25%; **(c)** 50%; **(d)** 70%.

### 3.4. January–March 2022 (Omicron variant)

For January–March 2022, we estimated value with consideration of vaccination and some restrictions on mass gathering (light and strict). The determination of light and strong restrictions can be found in Section 2.2.3.

At the beginning of the 2022 year, the Omicron variant has already spread around the globe and caused new COVID-19 outbreaks. However, during January–March 2022 period, Taiwan demonstrated the capability to control Omicron’s outbreak. The maximum number of local cases amounted to 82 that happened at the end of January 2022, and the average value of new local cases was around 20 cases per day (Figure 8).

**Figure 8.**
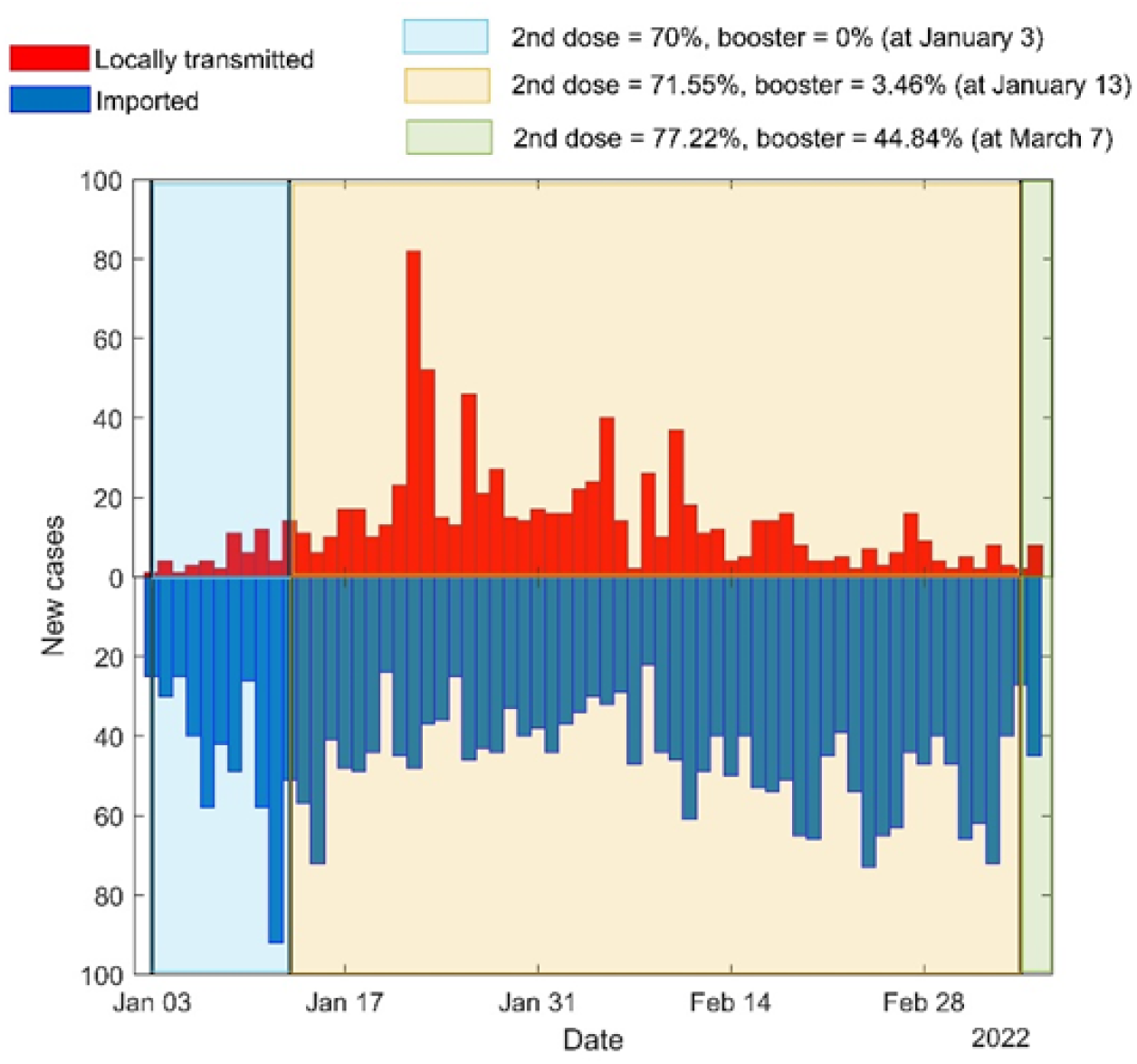
Official number of local and imported cases during January–March 2022 with proportion of vaccinated people at certain dates.

Estimation of was made for the beginning of January 2022 and for the beginning of March 2022, to show how a high level of vaccination together with other non-pharmaceutical interventions can control the outbreak (Figure 9). For the beginning of January 2022, vaccinated people by two doses were taken as 70% and by booster dose were taken as 0%. Since January 13, 71.55% is vaccinated with second dose and 3.46% booster [19].

**Figure 9.**
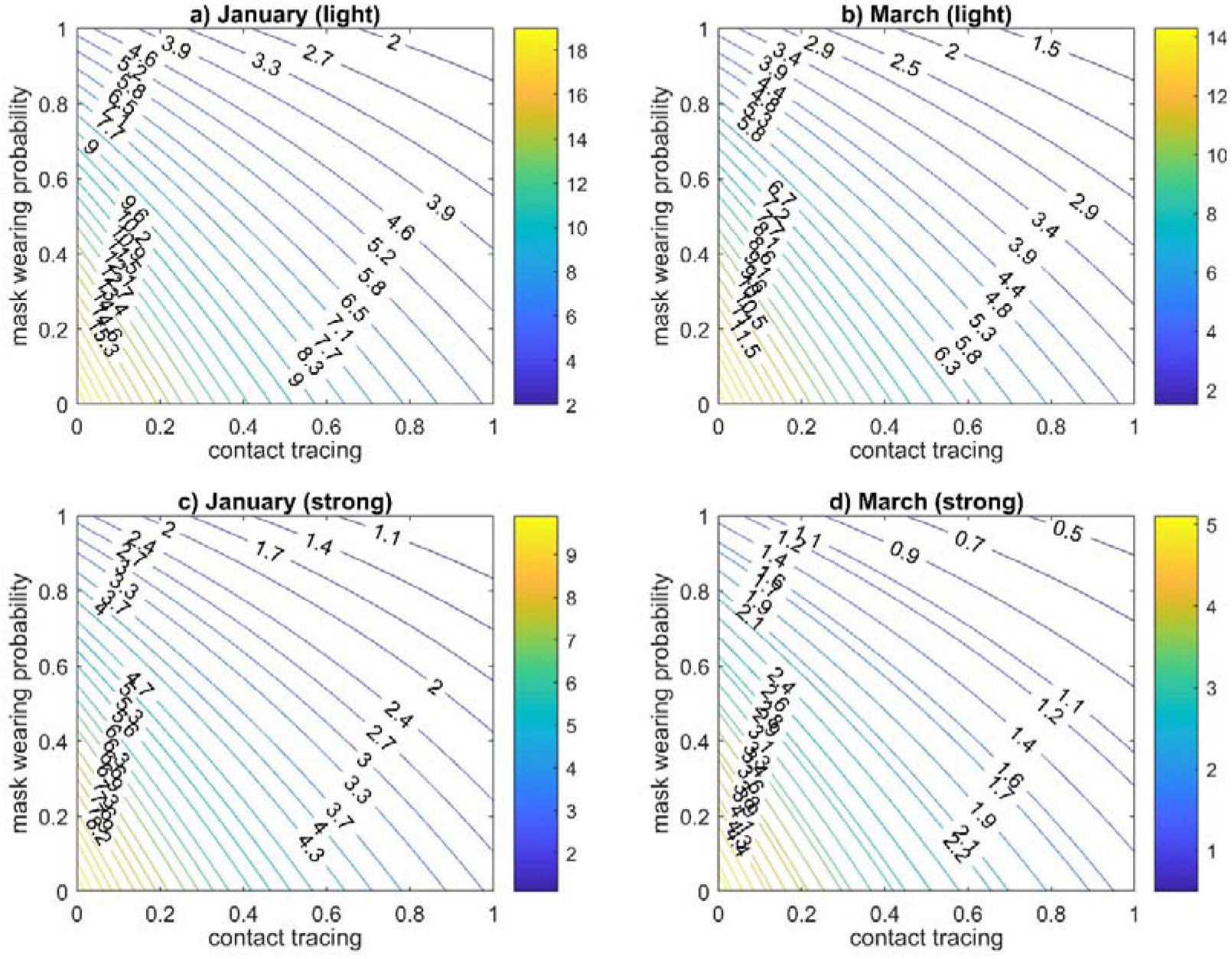
The basic reproduction number (R_0_) estimation for January–March 2022 period (with consideration of asymptomatic cases): **(a–b)** January–March 2022 with small gathering restrictions; **(c–d)** January–March 2022 with strong gathering restrictions.

Figures 9a and 9c represent the *R*_0_ estimation at the beginning of January for the different gathering restrictions efficiency: light restrictions with 15% efficiency (Figure 9a) and strong restrictions with 46% efficiency (Figure 9c). It can be seen that even with high proportion of mask-wearing (90%) and contact tracing (80–90%), *R*_0_ is equal to or even higher than 1. In Figure 9a, for *p =* 0.9 and *tr =* 0.9, *R*_0_ = 2.039. In Figure 9c, for *p =* 0.9 and *tr =* 0.9, *R*_0_ = 1.058. The obtained estimated values of *R*_0_ can explain the growth in number of new local cases at the end of January (Figure 8).

Figures 9b and 9d shows estimation of *R*_0_ values at the moment of March 7, 2022. Since the percentage of vaccinated people had been increased, especially vaccinated by booster dose (77.22% by two doses and 44.84% by booster) [17,19], this impacted the reduction of the *R*_0_ values. In Figure 9b, for *p =* 0.9 and *tr =* 0.9, *R*_0_ = 1.529. In Figure 9d, for *p =* 0.9 and *tr =* 0.9, *R*_0_ = 0.548. The obtained results of *R*_0_ during January–March 2022 can help to understand, how Taiwan was able to control the outbreak of the Omicron variant during this time. In addition, Figure 9 demonstrates that any small relaxations can lead to an increase in cases number.

## 4. Discussion and Conclusions

The results of our simulation of the spread of COVID-19 in Taiwan during the 2020–2022 years demonstrate that the improvement of a contact tracing program, the rapid implementation of measures to isolate infected individuals and restrict mass gathering, the wearing of surgical masks by 90% of the population, and fast increasing of vaccinated people can help to eliminate the local spread of the virus. However, it should be noted that only with the stable and careful implementation of all these interventions the estimated *R*_0_ can drop below one, which indicates the prevention of the spread of the virus. For example, even in areas with few confirmed cases, people should continue to wear masks not only where it is mandatory but in any public place. The results also indicate that for the outbreak control over new appearing COVID-19 variant, the interventions that were not considered during the previous COVID-19 variant, need to be involved. Moreover, to control outbreaks of the Delta or Omicron variants without very strict mass gathering restrictions, it is necessary to consistently increase the proportion of the vaccinated population. The vaccination program should also be improved with time, for example, by including booster dose or revaccination. In this work, we examined the handling of COVID-19 in Taiwan during 2020–2021 years to determine the best practices to help controlling of the virus spread before pharmaceutical interventions (vaccines) are available. Contact tracing, the wearing of masks, fast time of isolation, and mass gathering restrictions are effective tools but the majority of a population must be on board. This includes residents, who must observe measures and consistently wear a mask, as well as the government, which should introduce and facilitate transmission control measures and develop further strategies to address the pandemic.

In addition, in the current study, the estimation of the *R*_0_ was made for Taiwan during the January–March of the 2022 year. For this period, the vaccination has been already started to be one of the most important interventions such as mask-wearing and contact tracing. Only for three months (from January to March), the number of people vaccinated by a booster dose has been increased by more than 40% [17,19].

Universal access to masks and contact tracing to control transmission from symptomatic and asymptomatic individuals may reduce the number of COVID-19 infections. This is important since drastic interventions such as the lockdown of cities or countries or social distancing, while proven relatively effective to control the spread of COVID-19, are unsustainable in the long-term due to high social and economic costs. One recent study gives strong support for maintaining mask-wearing until and after achieving vaccination since using face masks can be cost-effective and cost-saving [78]. The Omicron variant emergence and the possible new variants in the future that can be more transmissible and reduce vaccine efficiency only increases the value and efficiency of mask-wearing. When countries gradually return to normal life from lockdown, it is essential to understand the effectiveness of less intrusive, more sustainable interventions, such as wearing a mask, contact tracing program, and some mass gathering restrictions as measures of source control even in places where COVID-19 vaccines are being involved [79].

Several early COVID-19 modeling studies examined the effect of lockdowns and physical distancing measures implemented in Wuhan, China. However, in 2020, several countries, including Taiwan, South Korea, Singapore, and Japan, have contained local transmission without drastic measures such as lockdowns and shelter-in-place orders, suggesting that if conventional transmission control measures are thoroughly implemented, an outbreak can be contained. At the beginning of 2022, Taiwan did not introduce strong mass gathering restrictions (as it was during May–July of 2021) but mask-wearing, contact tracing together with vaccination programs try to be kept at a high implementation level.

At the end of March 2022, Taiwan started to change its “zero COVID-19” policy to the new strategy. Some restrictions started to relax from March 2022. The new COVID-19 policy is no longer focused on the total termination of the outbreak but it is still oriented to encourage people for taking booster dose, to save the public’s ability to remain watchful, and take necessary measures to protect their health [80]. The new Taiwanese model is also oriented to reduce the burden on the medical system and maintain a normal life [80,81]. The priorities shift was caused by the fact that most of the cases (99.6%) in the 2022 year have mild or no symptoms [81].

Despite this new COVID-19 policy, Taiwan still continues to maintain mandatory mask-wearing, and, as of 11 April 2022, around 80% of the population had two vaccine doses while more than 50% had the booster dose [17,19].

Our findings are relevant for health authorities because they provide quantitative estimates of the effectiveness of both strict and light interventions. Continuation of the mandatory mask-wearing in Taiwan is consistent with our obtained results. Despite the vaccination, non-pharmaceutical interventions such as mask-wearing and contact tracing can play an important role. However, the effect of universal access to face masks as well as of contact tracing programs varies by country because of the highly individual nature of population behavior and recent country experience of struggle with some viruses.

## 5. Limitations

This study has some limitations. We used some assumptions for different parameter values since few parameters are very difficult to estimate. While there has been no forced lockdown, mandated social distancing, or obligatory cancellation of events in Taiwan, people have taken many voluntary protective measures due to fear of infection. It is uncertain whether, when life returns to normal and people become less cautious and fearful, mask-wearing will remain effective.

## Data Availability

All data produced in the present work are contained in the manuscript

## Acknowledgments

This research was funded by the National Health Research Institutes, Taiwan, grant number BN-109-PP-08, and the Ministry of Science and Technology, Taiwan, grant number MOST 106 2115 M 400 001. The funder has no role in study design, data collection and analysis, decision to publish, or preparation of the manuscript.

