## Supplementary for "The impact of surgical mask-wearing, contact tracing program, and vaccination on COVID-19 transmission in Taiwan from January 2020 to March 2022: a modelling study"

**for**

^†^Equal contributors

1. **Derivation of formulas for number of infected and non-infected passengers from Wuhan to Taiwan**

To estimate the number of passengers from Wuhan to Taiwan during January 2020, we model the number of infected people in Wuhan using the SEIR model [1]. Since we are interested only in the number of infected people, we use the first three equations of the model from [1]:

$\dot{S}_{W}=-\frac{S_{W}}{N_{W}}\left( \frac{R_{0}}{T_{inf}}I_{w}+z \right)+P_{IW}+P_{CW}-(\frac{P_{WI}+P_{WC}}{N_{W}})S_{W}$	(1)

$\dot{E}_{W}=\frac{S_{W}}{N_{W}}\left( \frac{R_{0}}{T_{inf}}I_{w}+z \right)-\frac{E_{w}}{T_{inc}}-(\frac{P_{WI}+P_{WC}}{N_{W}})E_{w}$	(2)

$\dot{I}_{W}=\frac{E_{w}}{T_{inc}}-\frac{I_{w}}{T_{inf}}-(\frac{P_{WI}+P_{WC}}{N_{W}})I_{w}$	(3)

where $S_{W}$, $E_{W}$, and $I_{W}$ are the number of susceptible, exposed, and infectious individuals, respectively, in the Wuhan region, with total population $N_{W}$. $T_{inc}$ and $T_{inf}$ are the mean incubation period and the infectious period respectively. $T_{s}$ represents the mean serial interval [2]. $R_{0}$ is the basic reproduction number for Wuhan model during this period; and $z$ is the zoonotic force of infection, equal to $86$ cases per day before Wuhan’s wet market closure on January 1, 2020, and $0$ thereafter. Based on the data from [1], which used 2019 Official Airline Guide (OAG) data for cities outside mainland China, excluding Hong Kong, the average daily number of international outbound air passengers in Greater Wuhan was $P_{WI}=3633$. The average daily number of international inbound air passengers for the same area was $P_{IW}=3546$ from January to February, 2019; the average number of domestic travelers in China from Wuhan $P_{WC}=502 013$ and to Wuhan $P_{CW}=487 310$ before 10 January (*Chunyun spring* festival on January 10); during the 40-day *Chunyun period*, the corresponding numbers were $P_{WC}=717 226$ and $P_{CW}=810 500$ [1]. In this system, as shown in Equations (1)–(3), the total number of infected people at time $t$ is $E_{W}(t)+I_{W}(t)$. A total of $2 363$ people flew from Wuhan to Taiwan in January 2020 [3]. Since Wuhan was closed to travel after January 23 2020 [4], we can assume that about $2396/23=104$ people travelled from Wuhan to Taiwan each day from January 1 to January 23. Thus, since we know the total number of cases in Wuhan for each day, we can estimate the proportion of infected $P_{WT}^{inf}$ and non-infected $P_{WT}$ passengers from Wuhan to Taiwan:

$P_{WT}^{inf}(t) = (E_{W}(t)+I_{W}(t))\cdot2363 /(23\cdot N_{W})$	(4)

$P_{WT}(t) = 23633/23 - P_{WT}^{inf}(t)$	(5)

This model is run together with the main model for Taiwan (Equations (27)-(38) in the paper) from 25 December, 2019, and the initial values, which are not equal to zero, are following:

$$S_{W}\left( 0 \right)=N_{W}-I_{W}\left( 0 \right); I_{W}\left( 0 \right)=86; N_{W}=19\cdot{10}^{6}.$$

The values of the parameters, used in the model (1)–(3), are

$$T_{inc}=5.75; T_{s}=7.5; T_{inf}=T_{s}-T_{inc}=1.75; R_{0}=2.68.$$

2. **The basic reproduction number estimation by the next-generation matrix (NGM)**

The model, which we investigate for the estimation of the basic reproduction number $R_{0}$, has the following form:

$\dot{S}_{n}=-\frac{\beta}{N}S_{n}X(1-tr)$	(6)

$\dot{S}_{m}=-\frac{\beta r}{N}S_{m}X\left( 1-tr \right)$	(7)

$\dot{A}_{n}=\frac{\alpha\beta}{N}S_{n}X\left( 1-tr\cdot{pr}_{a}^{s} \right)-\gamma_{1}A_{n}-\frac{tr\cdot{pr}_{a}^{s}}{T_{iso}}A_{n}$	(8)

$\dot{A}_{m}=\frac{\alpha\beta r}{N}S_{m}X\left( 1-tr\cdot{pr}_{a}^{s} \right)-\gamma_{1}A_{m}-\frac{tr\cdot{pr}_{a}^{s}}{T_{iso}}A_{m}$	(9)

$\dot{E}_{n}=\frac{(1-\alpha)\beta}{N}S_{n}X\left( 1-tr\cdot{pr}_{s}^{s} \right)-\frac{E_{n}}{T_{inc}}-\frac{tr\cdot{pr}_{s}^{s}}{T_{iso}}E_{n}$	(10)

$\dot{E}_{m}=\frac{(1-\alpha)\beta r}{N}S_{m}X\left( 1-tr\cdot{pr}_{s}^{s} \right)-\frac{E_{m}}{T_{inc}}-\frac{tr\cdot{pr}_{s}^{s}}{T_{iso}}E_{m}$	(11)

$\dot{I}_{n}=E_{n}/T_{inc}-I_{n}/T_{iso}$	(12)

$\dot{I}_{m}=E_{m}/T_{inc}-I_{m}/T_{iso}$	(13)

$\dot{C_{a}}=\frac{tr\cdot{pr}_{a}^{s}}{T_{iso}}(A_{n}+A_{m})-\gamma_{3}T_{a}$	(14)

$\dot{C_{s}}=\frac{tr\cdot{pr}_{s}^{s}}{T_{iso}}(E_{n}+E_{m})-\gamma_{4}T_{s}$	(15)

$\dot{Q}=(I_{n}+I_{m})/T_{iso}+\gamma_{4}T_{s}-\gamma_{2}Q$	(16)

$\dot{R}=(\gamma_{1}A_{n}+\gamma_{1}A_{m}+\gamma_{2}Q+\gamma_{3}T_{a})\left( 1-\delta\right)$	(17)

where $X={\mu A}_{n}+r\mu A_{m}+{\varepsilon E}_{n}+r{\varepsilon E}_{m}+I_{n}+rI_{m}$.

We want to mention, that here we did not include in the model the travelers from Wuhan.

Diekmann et al. proposed to define the basic reproduction number $R_{0}$ by using so-called the next-generation matrix (NGM) [5,6]. Using notation from their work [6], we denote the NGM by $\boldsymbol{K}$. Diekmann et al. determined $R_{0}$ as the dominant eigenvalue of $\boldsymbol{K}$ [5,6].

The NGM consists of two matrices: $\boldsymbol{T}$ is the transmission matrix, and $\boldsymbol{\Sigma}$ is the transition matrix. The sum $\boldsymbol{T}+\boldsymbol{\Sigma}$ describes the Jacobian matrix of the original nonlinear ODE system. The transmission matrix $\boldsymbol{T}$ determines the production of new infected people, the transition matrix $\boldsymbol{\Sigma}$ describes the changes inside the certain state.

To estimate $R_{0}$ by the NGM method, we use the system (6)–(17). This system has six infected states ($A_{n}$, $A_{m}$, $E_{n}$, $E_{m}$, $I_{n}$, $I_{m}$) and also six uninfected state ($S_{n}$, $S_{m}$, $C_{a}$, $C_{s}$, $Q$, $R$). The size of the population, which is not wearing of masks, is $S_{n}=\left( 1-p \right)N$ and the size of the population, which wear masks, is $S_{m}=pN$. The infected subsystem (8)–(13) is used in order to estimate the basic reproduction number. Setting $\boldsymbol{x}=\left[ A_{n}, A_{m}, E_{n}, E_{m}, I_{n}, I_{m} \right]^{T}$, we can write our linearized subsystem in the following form:

$\dot{\boldsymbol{x}}=(\boldsymbol{T}+\boldsymbol{\Sigma})\boldsymbol{x}$	(18)

$$\boldsymbol{T}=\left[ \begin{matrix} \mu t_{An} & \mu rt_{An} & \varepsilon t_{An} & \varepsilon rt_{An} & t_{An} & rt_{An} \\ \mu t_{Am} & \mu rt_{Am} & \varepsilon t_{Am} & \varepsilon rt_{Am} & t_{Am} & rt_{Am} \\ \mu t_{En} & \mu rt_{En} & \varepsilon t_{En} & \varepsilon rt_{En} & t_{En} & rt_{En} \\ \mu t_{Em} & \mu rt_{Em} & \varepsilon t_{Em} & \varepsilon rt_{Em} & t_{Em} & rt_{Em} \\ 0 & 0 & 0 & 0 & 0 & 0 \\ 0 & 0 & 0 & 0 & 0 & 0 \end{matrix} \right]$$

$$\boldsymbol{\Sigma}=\left[ \begin{matrix} -\gamma_{1}-\frac{tr\cdot{pr}_{a}^{s}}{T_{iso}} & 0 & 0 & 0 & 0 & 0 \\ 0 & -\gamma_{1}-\frac{tr\cdot{pr}_{a}^{s}}{T_{iso}} & 0 & 0 & 0 & 0 \\ 0 & 0 & -\frac{1}{T_{inc}}-\frac{tr\cdot{pr}_{s}^{s}}{T_{iso}} & 0 & 0 & 0 \\ 0 & 0 & 0 & -\frac{1}{T_{inc}}-\frac{tr\cdot{pr}_{s}^{s}}{T_{iso}} & 0 & 0 \\ 0 & 0 & \frac{1}{T_{inc}} & 0 & -\frac{1}{T_{iso}} & 0 \\ 0 & 0 & 0 & \frac{1}{T_{inc}} & 0 & -\frac{1}{T_{iso}} \end{matrix} \right]$$

where

$$t_{An}=\alpha\beta\left( 1-p \right)\left( 1-tr\cdot{pr}_{a}^{s} \right), t_{Am}=\alpha\beta rp\left( 1-tr\cdot{pr}_{a}^{s} \right),$$

$$t_{En}=\left( 1-\alpha\right)\beta\left( 1-p \right)\left( 1-tr\cdot{pr}_{s}^{s} \right), t_{Em}=\left( 1-\alpha\right)\beta rp\left( 1-tr\cdot{pr}_{s}^{s} \right),$$

$$\gamma_{1}=\frac{1}{T_{a,inf}}.$$

The matrix $\boldsymbol{T}$ has zero rows. We create an additional matrix $\boldsymbol{E}$ which consists of unit column vectors relating to only non-zero rows of $\boldsymbol{T}$ [6]:

$$\boldsymbol{E}=\left[ \begin{matrix} 1 & 0 & 0 & 0 \\ 0 & 1 & 0 & 0 \\ 0 & 0 & 1 & 0 \\ 0 & 0 & 0 & 1 \\ 0 & 0 & 0 & 0 \\ 0 & 0 & 0 & 0 \end{matrix} \right]$$

According to the study [6], the NGM can be found by matrix multiplication:

$\boldsymbol{K}=- \boldsymbol{E}^{T}\boldsymbol{T}\boldsymbol{\Sigma}^{-1}\boldsymbol{E}$	(19)

For our model (6)–(17) the NGM has the following form:

$$\boldsymbol{K}=\left[ \begin{matrix} K_{11} & K_{12} & K_{13} & K_{14} \\ K_{21} & K_{22} & K_{23} & K_{24} \\ K_{31} & K_{32} & K_{33} & K_{34} \\ K_{41} & K_{42} & K_{43} & K_{44} \end{matrix} \right]$$

where

$$K_{11}=T_{a,inf}T_{iso}\mu t_{An}/(T_{iso}+T_{a,inf}tr\cdot{pr}_{a}^{s}),$$

$$K_{12}=T_{a,inf}T_{iso}\mu rt_{An}/(T_{iso}+T_{a,inf}tr\cdot{pr}_{a}^{s}),$$

$$K_{13}=T_{iso}t_{An}(T_{iso}+\varepsilon T_{inc})/(T_{iso}+T_{inc}tr\cdot{pr}_{s}^{s}),$$

$$K_{14}=T_{iso}rt_{An}(T_{iso}+\varepsilon T_{inc})/(T_{iso}+T_{inc}tr\cdot{pr}_{s}^{s}),$$

$$K_{21}=T_{a,inf}T_{iso}\mu t_{Am}/(T_{iso}+T_{a,inf}tr\cdot{pr}_{a}^{s}),$$

$$K_{22}=T_{a,inf}T_{iso}\mu rt_{Am}/(T_{iso}+T_{a,inf}tr\cdot{pr}_{a}^{s}),$$

$$K_{23}=T_{iso}t_{Am}(T_{iso}+\varepsilon T_{inc})/(T_{iso}+T_{inc}tr\cdot{pr}_{s}^{s}),$$

$$K_{24}=T_{iso}rt_{Am}(T_{iso}+\varepsilon T_{inc})/(T_{iso}+T_{inc}tr\cdot{pr}_{s}^{s}),$$

$$K_{31}=T_{a,inf}T_{iso}\mu t_{En}/(T_{iso}+T_{a,inf}tr\cdot{pr}_{a}^{s}),$$

$$K_{32}=T_{a,inf}T_{iso}\mu rt_{En}/(T_{iso}+T_{a,inf}tr\cdot{pr}_{a}^{s}),$$

$$K_{33}=T_{iso}t_{En}(T_{iso}+\varepsilon T_{inc})/(T_{iso}+T_{inc}tr\cdot{pr}_{s}^{s}),$$

$$K_{34}=T_{iso}rt_{En}(T_{iso}+\varepsilon T_{inc})/(T_{iso}+T_{inc}tr\cdot{pr}_{s}^{s}),$$

$$K_{41}=T_{a,inf}T_{iso}\mu t_{Em}/(T_{iso}+T_{a,inf}tr\cdot{pr}_{a}^{s}),$$

$$K_{42}=T_{a,inf}T_{iso}\mu rt_{Em}/(T_{iso}+T_{a,inf}tr\cdot{pr}_{a}^{s}),$$

$$K_{33}=T_{iso}t_{Em}(T_{iso}+\varepsilon T_{inc})/(T_{iso}+T_{inc}tr\cdot{pr}_{s}^{s}),$$

$$K_{44}=T_{iso}rt_{Em}(T_{iso}+\varepsilon T_{inc})/(T_{iso}+T_{inc}tr\cdot{pr}_{s}^{s}),$$

and $R_{0}$ is the dominant eigenvalue of matrix $\boldsymbol{K}$:

$R_{0}=(X_{1}\left( T_{iso}^{3}+\varepsilon T_{iso}^{2}T_{inc} \right)+T_{iso}^{2}T_{a,inf}X_{2}+T_{a,inf}T_{inc}T_{iso}X_{3})/(T_{iso}^{2}+T_{inc}T_{iso}tr\cdot{pr}_{s}^{s}+T_{a,inf}T_{iso}tr\cdot{pr}_{a}^{s}+T_{a,inf}T_{inc}{tr}^{2}{pr}_{a}^{s}{pr}_{s}^{s})$	(20)

where

$$X_{1}=t_{En}+rt_{Em},$$

$$X_{2}=\mu t_{An}+\mu rt_{Am}+t_{En}tr\cdot{pr}_{a}^{s}+rt_{Em}tr\cdot{pr}_{a}^{s},$$

$$X_{3}=\varepsilon t_{En}tr\cdot{pr}_{a}^{s}+\mu t_{An}tr\cdot{pr}_{s}^{s}+\varepsilon rt_{Em}tr\cdot{pr}_{a}^{s}+\mu rt_{Am}tr\cdot{pr}_{s}^{s}.$$

Equation (20) is the estimation of the basic reproduction number $R_{0}$ with consideration of the asymptomatic cases.

In the current study, we also use the NGM approach to evaluate the basic reproduction number without taking into account the asymptomatic cases. For this instance, only equations (10)–(13) are used as infected subsystem. The other components of the NGM method change as follows:

$\boldsymbol{x}=\left[ E_{n}, E_{m}, I_{n}, I_{m} \right]^{T},$	(21)

$\boldsymbol{T}=\left[ \begin{matrix} \varepsilon t_{En} & \varepsilon rt_{En} & t_{En} & rt_{En} \\ \varepsilon t_{Em} & \varepsilon rt_{Em} & t_{Em} & rt_{Em} \\ 0 & 0 & 0 & 0 \\ 0 & 0 & 0 & 0 \end{matrix} \right]$	(22)

$\boldsymbol{\Sigma}\boldsymbol{=}\left[ \begin{matrix} \boldsymbol{-}\frac{\boldsymbol{1}}{\boldsymbol{T}_{\boldsymbol{inc}}}\boldsymbol{-}\frac{\boldsymbol{tr\cdot}\boldsymbol{pr}_{\boldsymbol{s}}^{\boldsymbol{s}}}{\boldsymbol{T}_{\boldsymbol{iso}}} & \boldsymbol{0} & \boldsymbol{0} & \boldsymbol{0} \\ \boldsymbol{0} & \boldsymbol{-}\frac{\boldsymbol{1}}{\boldsymbol{T}_{\boldsymbol{inc}}}\boldsymbol{-}\frac{\boldsymbol{tr\cdot}\boldsymbol{pr}_{\boldsymbol{s}}^{\boldsymbol{s}}}{\boldsymbol{T}_{\boldsymbol{iso}}} & \boldsymbol{0} & \boldsymbol{0} \\ \frac{\boldsymbol{1}}{\boldsymbol{T}_{\boldsymbol{inc}}} & \boldsymbol{0} & \boldsymbol{-}\frac{\boldsymbol{1}}{\boldsymbol{T}_{\boldsymbol{iso}}} & \boldsymbol{0} \\ \boldsymbol{0} & \frac{\boldsymbol{1}}{\boldsymbol{T}_{\boldsymbol{inc}}} & \boldsymbol{0} & \boldsymbol{-}\frac{\boldsymbol{1}}{\boldsymbol{T}_{\boldsymbol{iso}}} \end{matrix} \right]$	(23)

$\boldsymbol{E}=\left[ \begin{matrix} 1 & 0 \\ 0 & 1 \\ 0 & 0 \\ 0 & 0 \end{matrix} \right]$	(24)

The NGM is found by the formula (19). For the model (6)–(17) but with only symptomatic cases the NGM has the following form:

$$\boldsymbol{K}=\left[ \begin{matrix} K_{11} & K_{12} \\ K_{21} & K_{22} \end{matrix} \right]$$

where

$$K_{11}=T_{iso}t_{En}(T_{iso}+\varepsilon T_{inc})/(T_{iso}+T_{inc}tr\cdot{pr}_{s}^{s}),$$

$$K_{12}=T_{iso}rt_{En}(T_{iso}+\varepsilon T_{inc})/(T_{iso}+T_{inc}tr\cdot{pr}_{s}^{s}),$$

$$K_{21}=T_{iso}t_{Em}(T_{iso}+\varepsilon T_{inc})/(T_{iso}+T_{inc}tr\cdot{pr}_{s}^{s}),$$

$$K_{22}=T_{iso}rt_{Em}(T_{iso}+\varepsilon T_{inc})/(T_{iso}+T_{inc}tr\cdot{pr}_{s}^{s}),$$

and $R_{0}$ (for model with only symptomatic cases consideration) is also the dominant eigenvalue of matrix $\boldsymbol{K}$:

$R_{0}=\left( T_{iso}^{2}+\varepsilon T_{iso}T_{inc} \right)\left( t_{En}+rt_{Em} \right)/(T_{iso}+T_{inc}tr\cdot{pr}_{s}^{s}).$	(24)
